# An *R*_*t*_- based model for predicting multiple epidemic waves in a heterogeneous population

**DOI:** 10.1101/2022.10.27.22281524

**Authors:** Razvan Romanescu, Songdi Hu, Douglas Nanton, Mahmoud Torabi, Olivier Tremblay-Savard, Md Ashiqul Haque

## Abstract

Relaxing the homogeneous mixing assumption in a population is often necessary to improve fits of epidemic models to observed infection counts. Establishing a link between observed infections and the underlying network of contacts is paramount to understanding how the network structure affects the speed of spread of a pathogen. In this paper we argue that introducing a flexible structure for the effective reproductive number (Rt) over the course of an epidemic allows for a more realistic description of the network of social contacts. This, in turn, produces better retrospective fits, as well as more accurate prospective predictions of observed epidemic curves. We extend this framework to fit multi-wave epidemics, and to accommodate public health restrictions on mobility. We demonstrate the performance of this model by doing a prediction study over two years of the SARS-CoV2 pandemic.

## 1. Background

Since the introduction of the susceptible-infected-removed (SIR) model of epidemic spread nearly a century ago (Kermack & McKendrick, 1927), the homogeneity of individuals in terms of their chance of interacting with one another has been a mainstay assumption in infectious disease models to this day. This is despite ample evidence that some individuals contribute much more to disease transmission than others (Wang et al, 2020), and that the assumption of homogeneity, also called mass action, produces poor fits to real data (Stack et al, 2012; Romanescu & Deardon, 2017). We identify the following two reasons why the mass action assumption continues to be used in practice. First, a homogeneous population leads to simple mathematical ordinary differential equation (ODE) models that have a physical analog in the time progression of chemical reactions (where the mass action principle also originates). Second is the fact that a heterogeneous models of spread requires additional assumptions about how individuals differ in their ability to transmit disease. This extra information may not be easily available, or if it is, it can increase the complexity or computational burden of the models far beyond simple ODEs (such as, for instance, agent-based models e.g., Deardon et al., 2010).

Network models are one alternative to mass action models. They describe the population via a network with individuals as nodes and contacts as edges, and model individual heterogeneity via a different number of infectious contacts for each individual. A network with first order properties only is characterized by the distribution of a node’s degree (the number of edges emanating from it). Higher order properties relate to community structures beyond each node’s own contacts. We will restrict our attention to first order properties, as this provides a rich enough family of models to work with. First analyzed mathematically in Newman (2003), epidemics over networks have some unique properties, for instance an epidemic can occur for arbitrarily low transmissibility, provided that the degree distribution is skewed enough (heavy tailed). In practice, network models are adept at explaining patterns in real data that cannot be accounted for by mass action models, such as sharp peaks of the epidemic curve, and an initial growth rate that far exceeds that predicted using textbook values of the basic reproductive number (*R*_0_). Note that with a mass action assumption, the number of new infections arising from one infected (the effective reproductive number, *R*_*t*_) starts from *R*_0_ and decays linearly with the remaining susceptible fraction, *R*_*t*_ = *R*_0_*S*_*t*_, as *S*_*t*_ progresses from 1 to *S*_∞_ < 1.

While network models offer much flexibility to explain observed data, they have the obvious disadvantage of requiring knowledge of the underlying network, at least as the probability distribution of contacts per individual. This is difficult to observe, and although it has been attempted in limited contexts (staff in a healthcare setting, see e.g., Hornbeck et al., 2012; Machens et al., 2013), these don’t generalize well to the entire population. Fortunately, the flexibility of network models (at least at the first degree) can be entirely captured via the shape of decline in *R*_*t*_ over the course of the epidemic, as we will argue in this paper. Thus, modeling efforts for predicting the epidemic curve can be focused entirely on estimating *R*_*t*_, and this will unlock the benefits of network models without needing to reference the underlying network. The idea that a flexible form for *R*_*t*_ can offer realism to an SIR model is not new. It has been considered in various incarnations before: as a ‘generalized rate of infection’ modification to traditional ODE models (Stroud et al., 2006; Connell et al., 2009); or *R*_*t*_ as a decreasing function of time in Chowell et al. (2016) and Wu et al. (2020). This paper takes a novel approach in that it starts by assuming the existence of an underlying network, and derives formulas for *R*_*t*_ that are consistent with diffusion processes over this network. The significance is that there are normative rules for how *R*_*t*_ progresses, especially in the context of loss of immunity and restrictions on the contact network. Although modelers have addressed these phenomena using ad-hoc techniques, having a theoretical framework in place provides a more solid foundation for understanding these complex processes, as well as may lead to improved prediction performance.

The paper is organized as follows: in Section 2 we present the method, including fitting to count data. In Section 3 we fit the model to COVID-19 infection counts from the state of New York and the city of Winnipeg and infer the *R*_*t*_ curve for that population using a couple of possible shapes. We also run a prediction study to demonstrate the utility of the *R*_*t*_ curves in predicting the future epidemic trajectory. We conclude with a discussion and possible directions for future research.

## 2. Methods

### 2.1 Prediction model

We formulate the prediction model in an SIR framework discretized at an appropriate time step (usually one day). We start from an estimator of the effective reproduction number (Nishiura & Chowell, 2009; Cori et al., 2013): 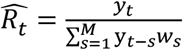, where *y*_*t*_ is the number of infections on day *t*; *w*_*s*_ is the generation interval, and more exactly the probability of the time between a primary and secondary infection events being *s* days; and *M* is the maximum range of the generation interval. Then we can re-express this formula to predict incidence at time *t* + 1 as in Abbott et al. (2020) and Nishiura & Chowell (2009):

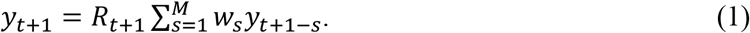

The susceptible fraction is updated as:

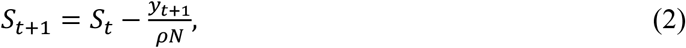

where N is the total population size and *ρ* is the underreporting rate (Romanescu & Deardon, 2017). Next, we turn our attention to predicting *R*_*t*+1_ in (1). Based on previous work from network-based epidemic models, we model the effective reproductive number as *R*_*t*+1_ = *αS*_*t*_*c*(*S*_*t*_), where *α* is the person to person probability of transmission and *c*(*S*_*t*_) is the average contact rate of the set of infectious individuals, which decays as *S*_*t*_ decreases (see the Appendix for a review of the theory).

We consider the shapes for *c*(·) given in <u>Table 1</u>. Since *α* is not estimable on its own from time series data, we parameterize *αc*(*S*_*t*_) as a whole, with parameters that can be estimated.

**Table 1.** Functional forms of the average contact rate (times α).

| Shape of decay | Parameters | $\alpha \times c(S_t)$ |
| --- | --- | --- |
| Exponential | $a_1, a_2, a_3$ | $a_0 \{e^{a_1(S_t - a_2)} - 1\}$ |
| Two piece exponential | $a_1, a_2, a_3, a_4$<br>($a_4 > a_2; a_3 > a_1$) | $e^{a_1(S_t - a_2)} + \max(e^{a_3(S_t - a_4)}, 1) - 2$ |
| Shifted inverse | $a_0, a_1$<br>($a_1 > 1$) | $\frac{a_0}{a_1 - S_t}$ |

**Table 2.**
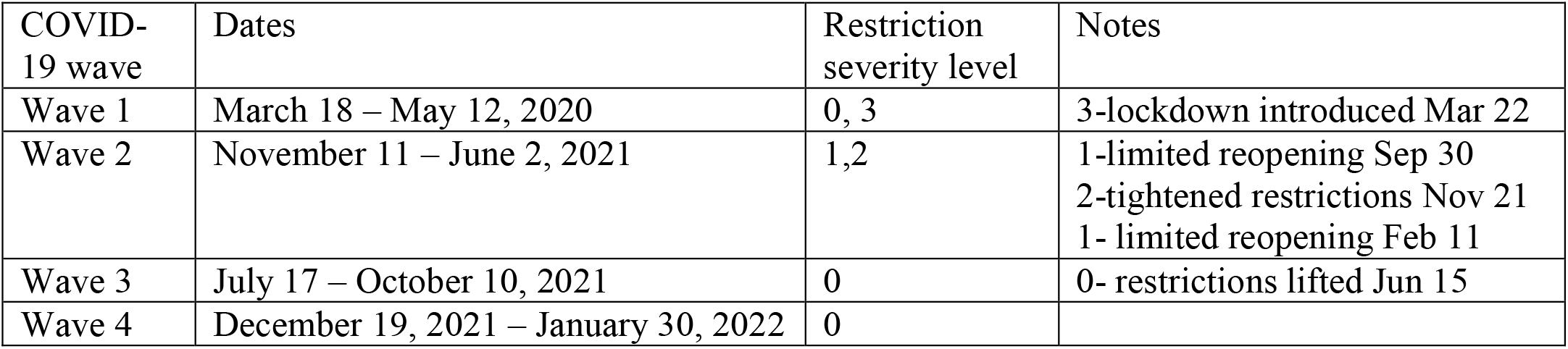
Wave definitions and severity level of restriction on the social network for the state of NY.

| COVID-19 wave | Dates | Restriction severity level | Notes |
| --- | --- | --- | --- |
| Wave 1 | March 18 – May 12, 2020 | 0, 3 | 3-lockdown introduced Mar 22 |
| Wave 2 | November 11 – June 2, 2021 | 1,2 | 1-limited reopening Sep 30<br>2-tightened restrictions Nov 21<br>1- limited reopening Feb 11 |
| Wave 3 | July 17 – October 10, 2021 | 0 | 0- restrictions lifted Jun 15 |
| Wave 4 | December 19, 2021 – January 30, 2022 | 0 |  |

Our choices of functional forms in Table 1 allow for significant flexibility in the shape of *R*_*t*_, and are tailored to accommodate dynamics consistent with network theory. In particular, after an initial stochastic phase following the entry of a new pathogen in a population (when there is a non-zero chance of the epidemic dying out), transmission enters the ‘mainstream’ community. At this time *R*_*t*_ immediately shoots up to its maximum value, as transmission is driven by superspreaders (i.e., the most connected individuals); then it starts decaying in a deterministic fashion, according to the structure of the network. The shapes we consider allow for potentially high values of *R*_*t*_ at the start of the epidemic, a varying degree of curvature during the decline, and an adjustable maximum size of the epidemic at *S*_*t*_ = *a*_2_ for the exponential shapes (the final size condition is that *R*_*t*_ drops to zero, which is similar to that in Miller et al., 2012). The two piece exponential is meant to accommodate a precipitous decay in *R*_*t*_ at rate *a*_3_ when the population is fully susceptible (*S*_*t*_ = 100%), for the first few percentage points drop in *S*_*t*_ until level *a*_4_; after which the decay proceeds at the much smaller rate *a*_1_. The shifted inverse is another parsimonious solution for producing both a high initial value as well as a kink in the shape of *R*_*t*_.

Together, equations (1 – 2) and the shape of *c*(·) define the epidemic model used to describe incidence within waves. We define the start of a wave to be the day of an observed maximum 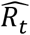; as mentioned above, spread dynamics tend to be stochastic before that time, so we do not try to make predictions prior to the maximum. The end of a wave will be the day of a minimum in 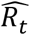, which should be well below one. Retrospectively, there should be no problem to identify these time points. Prospectively, however, we need a way to confirm a recent maximum or minimum. So, for the maximum we recommend waiting one day to confirm a high observed 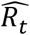 value as the true maximum. By ‘high’ we generally mean a value of around 2 or higher. For the minimum, we recommend waiting 2 weeks for confirmation. This asymmetry reflects the importance of timeliness in making predictions at the beginning of a wave, while at the end predictions will be small and less important, and the interest is more in ensuring that the wave has ended. Henceforth, we will use retrospectively defined waves, which is fair because here we are more concerned with the times we should start fitting the model, rather than pinpointing the exact start time of a new wave. In practice, the start of a new wave receives much attention and expertise from the broader epidemiological community; thus we make no claims that this definition of waves is the best, except perhaps to say that it makes the most sense to make predictions in that interval, at least from a network modeling perspective. A similar framework has been used for early identification of seasonal epidemic waves, and we refer the interested reader to some of our previous work (Romanescu & Deardon, 2019).

### 2.2. Including restrictions on the social network

In order to fit count data over longer periods of time, it is necessary to accommodate changing restriction measures and other public health interventions, as well as changing transmissibility due to mutations in the pathogen, vaccinations, etc. Our model can accommodate two types of regime shifts: those that affect the underlying network structure, such as workplace closures and limits on gatherings, and those that affect the person-to-person probability of transmission *α*. The latter category is fairly broad, and captures mask mandates, vaccinations, as well as any changes in the pathogen itself, and often combinations of these factors. Changes in person-to-person probability of transmission result in scaling *R*_*t*_ directly by the amount of relative change, and can be accommodated by adding a scale parameter which multiplies *R*_*t*_ in each wave beyond the first.

Regime shifts that affect the network structure are currently modeled in the literature as a drop in transmissibility across the board, similar to a change in transmissibility. Except in the homogeneous mixing case, this is incorrect over networks because restrictions on gatherings (or a lockdown) effectively reduce the fraction of individuals with many connections, while increasing the fraction with few connections. We will model this phenomenon by imposing an exponential tail to the degree distribution of the underlying network. This is equivalent to changing the average contact rate *c*(*S*_*t*_) in the expression of *R*_*t*_ to *c*(*ψS*_*t*_), for an appropriately chosen parameter *ψ* < 1 (see Appendix for proof). The intuitive explanation is that an epidemic that starts on a restricted network has similar dynamics in *c*(*S*_*t*_) as one that has progressed on an unrestricted network up to 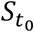 at time *t*_0_ and we are restarting the count of susceptibles 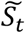 from 1 by now considering the entire population to be the susceptibles at time *t*_0_. The average contact rate will now be 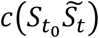, for *t* > *t*_0_ and the scaling factor *ψ* in this case is 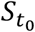. Essentially, restrictions will result in a lower contact rate of the infectious set, one normally associated to a later stage in the epidemic. The expression of *R*_*t*_ under such restriction becomes:

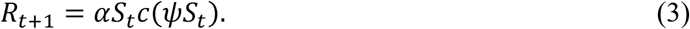

One may model less or more severe network restrictions using different *ψ* values at different times, with a lower value indicating more deleted connections. Parameters *ψ*_1_ > *ψ*_2_ > *ψ*_3_ > ⋯ corresponding to increasing severity levels 1,2,3, … are included in the estimation scheme described next. We assume a two week adjustment period following the introduction of a new restriction, during which *ψ* transitions linearly from the old to the new value.

### 2.3 Estimation procedure

First, denote by 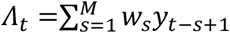, the denominator in the definition of *R*_*t*_ from Section 2.1. Following Romanescu & Deardon (2019), we can write the number of new infections as a sum of secondary infections from each infected individual counted in *Λ*_*t*_, thus: 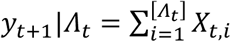, where [*Λ*_*t*_] is the closest integer to *Λ*_*t*_, and *X*_*t,i*_ are iid expansion factors, or counts of secondary infections for each infected individual. If we make the assumption that *Λ*_*t*_ is relatively stable, both because it’s a (weighted) sum, and also because in practice interest tends to be in short term predictions, meaning that some of the terms in *Λ*_*t*_ will tend to be based on the recent past, hence constants; then we can treat *Λ*_*t*_ as fixed, and we have the approximation *E*(*y*_*t*+1_|*Λ*_*t*_) = *Λ*_*t*_*E*(*X*_*t*_) and Var(*y*_*t*+1_|*Λ*_*t*_) = *Λ*_*t*_Var(*X*_*t*_). Importantly, the theoretical mean and variance of *X* have been derived previously (see Appendix for details), and although we don’t have information about the network degree distribution, the following formulation for the variance of *X* is reasonable: Var(*X*_*t*_) = *E*(*X*_*t*_)[*u* + *vE*(*X*_*t*_)], where *E*(*X*_*t*_) = *R*_*t*_, and *u, v* will be estimated from data.

Thus, the conditional count *y*_*t*+1_ works out to be:

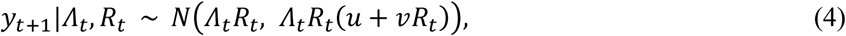

where normality is justified due to summing over a relatively large number of terms (*Λ*_*t*_), and also because *X*_*t*_ < *N* has finite moments. We can now proceed to fit the model parameters using all available case data via maximum likelihood estimation (MLE). The likelihood function for the parameter vector ***θ*** follows from distribution (4) as

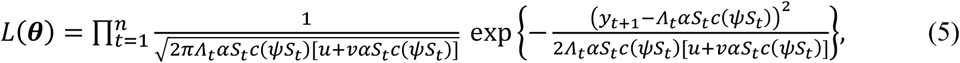

where (*y*_1_, *y*_2_, …, *y*_*n*_) are the daily observed new infections, and 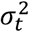 denotes the variance of the normal in Equation (4).

To find the MLE, one needs to maximize the log-likelihood over the parameter set, using all the data available at the present time. This is done using the “BFGS” routine of the optim function in R. Standard errors of the estimated parameters are computed using the large sample approximation to the MLE. Namely, the observed Fisher information matrix is obtained as the Hessian of the negative log-likelihood at the MLE, which is returned by the optimizer. This matrix is inverted to yield the variance-covariance matrix of the vector of estimates.

### 2.4. Extension to SIRS: accounting for loss of immunity

As the pathogen evolves, recovered individuals that had acquired natural immunity to infection may find themselves once again susceptible to new circulating strains. SIRS models describe the mechanism by which recovered individuals transition back to the susceptible compartment. Usually, an extra parameter, *ν*, defines the rate of flow per year between these two compartments, that is assumed to be constant through time. The complication with network models is that recovered individuals tend to be more well connected than individuals who were never infected, since they include the individuals who got infected early on in the epidemic. To model this, first denote by *t*_*l*_ and *T*_*l*_ the start and end times (in years) of epidemic wave *l*, so that *t*_1_ < *T*_1_ < *t*_2_ < *T*_2_ < ⋯. Notice that 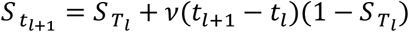, since at the start of the current wave (*l* + 1) the susceptible fraction is made up of individuals who remained susceptible at the end of the previous wave 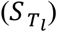 plus the newly susceptible who have lost their immunity somewhere in between *t*_*l*_ and *t*_*l*+1_ (these are a fraction of all preciously infected individuals 1 − 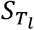). For simplicity we assume all individuals who lose their immunity do so at the start of a new wave.

We can demonstrate that the degree distribution of susceptible nodes at the start of a new wave (*t*_*l*+1_) is a mixture of the degree distribution of the susceptible set at the end of the previous wave (*T*_*l*_), and the original distribution, with weights 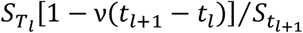 and 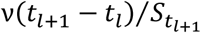, respectively (see Appendix). In terms of the transmission dynamics, this is conceptually similar to splitting the susceptible population into two subnetworks having different degree distributions and spreading infection at different rates.

We specify the following model in the SIRS case for each wave *l* ≥ 2 which defines different contact rates for each of the two subsets of the susceptible population:

(a) one subset of size 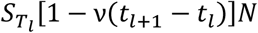 individuals has a contact rate (average number of contacts) of the infectious set which follows 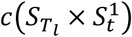, where 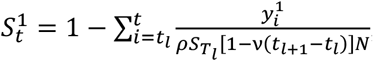, and 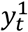 is the number of cases at *t* from this subset. This behavior is similar to starting the epidemic from fraction 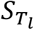 in the first season.

(b) for the remainder of the susceptible population (counting *v*(*t*_*l*+1_ − *t*_*l*_)*N* individuals at *t*_*l*+1_), the contact rate is modeled by 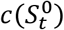, where 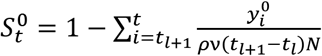, i.e., the same as starting in a fully susceptible population.

The following system of equations gives the number of new cases at the next time step, both in aggregate, and disaggregated by subset:

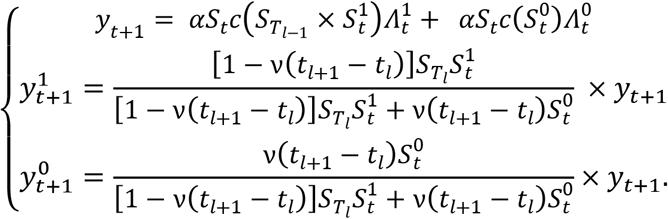

A justification of these assumptions is found in the Appendix. Thus the model computes recursively the aggregate and disaggregated number of new cases according to the following scheme:

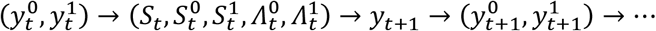

For fitting and prediction only the aggregated counts are used. The expression for *c* is as before, the only difference is the extra parameter *ν* which will be estimated as part of the likelihood function.

As a note on inference, there are two options for computing the loss function, in this case the likelihood, namely the sequential one-step-ahead inference, or the full curve approach. In the sequential approach, the likelihood (5) is computed by updating *S*_*t*_ and *Λ*_*t*_ with the actual (observed) values of *y*_*t*_. In the full curve approach, observed values are still used between waves, however, once a wave starts, model computed values 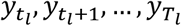, for all *l* are used to update *S*_*t*_ and *Λ*_*t*_ in the likelihood, as well as 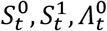, and 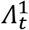. Both of these approaches can be used for retrospective model fitting, however, we use the full curve approach because this is the way ODE models are fit: for a set of parameters the full curves are produced, and then a loss function is computed between the actual epidemic curve and the model curve. This approach may also converge faster, as for parameter settings far from the MLE, errors accumulate faster than in the sequential approach, producing visibly divergent curves. One important advantage to mention over ODE models, though, is that this model is self-priming, meaning that it doesn’t require initial conditions to be estimated, hence reducing the number of necessary parameters to be fitted.

For predictions we should use all available information, and so it is necessary to use a combination of the previous approaches, namely actual values are used to update all quantities up to the present time, and the full curve approach is used to predict the future trajectory.

## 3. Results

### 3.1 Implementation details and settings

We fit the model to COVID-19 count data from two locations: the state of New York, and the city of Winnipeg. New York has been studied in our previous work on influenza (Romanescu & Deardon, 2017), and was found to be best described by relatively heavy-tailed distributions for the number of contacts of individuals. Although SARS-CoV2 and influenza are distinct respiratory diseases, with different transmissibility and serial interval profiles, based on our factorization (3) and update equation (1) the contact rate *c*(·) should be independent of these epidemiological factors. Winnipeg is a much smaller, provincial city in the Canadian Prairies, which stretches horizontally over a wide area, and where we would expect more homogeneity in transmission.

For the New York dataset (NY Times website), we define the epidemic waves based on the local maxima and minima of the empirical *R*_*t*_, as outlined in subsection 2.1. We identify four waves up to our cutoff of March 1^st^ 2022, as shown in Figure S1, in the Supplementary Material. The date ranges are shown in

Table 1. By searching through public health orders, we find restrictions with severities varying from 0 – 3 to have been introduced at the times listed in Table 1. For the Winnipeg data (Manitoba Health website), a similar table is given in the Supplementary Material (Table S1). For the analysis of both datasets, we assume the serial interval (*w*_*s*_) for COVID-19 from Nishiura et al. (2020) who model it as log-normal with median 4.0 days and standard deviation 2.9 days; we cap this at *M* = 20 days. The under-reporting rate (*ρ*) is informed from Irons & Raftery (2021) who compute the undercount fraction; this is 1/*ρ*, so we obtain *ρ* to be 1/2.1 or 47.6% for NY. There is no similar fraction computed for Winnipeg, so we use the value for North Dakota from the same source, due to geographical proximity; this is 1/1.7, or 58.8%. The population sizes for the state of NY and the Winnipeg regional health authority are taken from the US Census Bureau and from the Government of Manitoba, respectively.

### 3.2 Fit to the full dataset and implied *R*_*t*_ curve

We first fit our model to the full time series of infection counts, for all shapes in Table 1. The parameter estimates for each contact rate curve *c*(·) and the mass action model are given in Table 3. Note that for the mass action model, *ψ* should be interpreted as a constant multiplier to the transmissibility for the duration of the restriction. For the variance parameters *u* and *v* in formula (4), it was found that including both, or only one did not make a noticeable difference in the maximum likelihood, so we set *v* = 0 and only estimate *u*.

**Table 3.** Parameter estimates for NY state, using the full dataset. Maximum likelihood estimates and their standard deviations are shown. The (negative) log-likelihood value and the AIC are given at the bottom.

| Parameter |  | Definition | two-piece<br>exponential | one piece<br>exponential | shifted<br>inverse | mass<br>action |
| --- | --- | --- | --- | --- | --- | --- |
| $\alpha \times c(S_t)$ | $a_0$ | linear multiplier | – | 92.91<br>(10.46) | 0.54<br>(0.017) | 3.40<br>(0.046) |
| | $a_1$ | speed of decay | 1.26 (–) | 0.04<br>(0.043) | 1.11<br>(0.007) | – |
| | $a_2$ | final size | 0.00 (–) | 0.00 (–) | – | – |
| | $a_3$ | speed of decay (early) | 5.31<br>(0.041) | – | – | – |
| | $a_4$ | transition point/shift | 0.79(–) | – | – | – |
| $\nu$ | | loss of immunity rate | 1.55<br>(0.596) | 0.00<br>(0.003) | 1.10<br>(0.28) | 0.95<br>(-) |
| relative variant transmissibility<br>($\nu_1 = 1$) | $\nu_2$ | for second wave | 1.26<br>(0.031) | 0.46<br>(0.015) | 1.26<br>(0.040) | 1.33<br>(0.006) |
| | $\nu_3$ | for third wave | 0.26<br>(0.011) | 0.59<br>(0.019) | 0.31<br>(0.042) | 0.35 (-) |
| | $\nu_4$ | for fourth wave | 0.46<br>(0.043) | 1.06<br>(0.044) | 0.59<br>(0.082) | 0.41 (-) |
| restriction factors ($\psi$) | $\psi_1$ | limited reopening | 0.56 (-) | 1.00<br>(-) | 0.58<br>(0.014) | 0.24<br>(0.002) |
| | $\psi_2$ | tightened restrictions | 0.51 (-) | 0.90<br>(0.023) | 0.52<br>(0.014) | 0.24<br>(0.003) |
| | $\psi_3$ | during lockdown | 0.51 (-) | 0.43<br>(0.007) | 0.52<br>(0.017) | 0.24<br>(0.003) |
| $u$ | | overdispersion factor | 1140.53<br>(82.23) | 1652.72<br>(126.616) | 847.339<br>(63.268) | 2299.676<br>(110.415) |
| $-l(\theta)$ | | neg. log likelihood | 3615.605 | 3684.445 | 3577.093 | 3890.094 |
| # of parameters |  |  | 13 | 12 | 11 | 10 |
| AIC |  | Akaike Information Criterion | 7257.2 | 7392.9 | 7176.2 | 7800.2 |

The fitted curves are given in Figure 1. As we can see from the plot, the heterogeneous models (i.e., those with flexible *R*_*t*_) tend to fit observed data better than the mass action model, and this is confirmed by the AIC in Table 3. In particular, the mass action model has trouble accommodating sharp and early peaks. For the exponential curve models, it seems that the two piece exponential fits significantly better that the one piece, based on the AIC. In particular, it seems that the one piece model fits wave 3 poorly; the fits for the other waves are similar between the two models. The shifted inverse is the best fitting curve, suggesting that this shape is likely the most realistic to describe the decay in contact rate, at least for this dataset. We plot all the implied *R*_*t*_ curves in Figure 2. The differences between curves are quite large, and so are the implications for spread dynamics. The two-piece exponential is able to accommodate a higher *R*_*t*_ at the beginning of the epidemic (and this was the purpose for considering this shape). The curves also differ in the time they cross the endemic threshold; for example, if the two piece exponential is the correct model, then epidemics will peak at a smaller fraction of cumulative infections, compared to the mass action model, which peaks when many more individuals get infected. Overall, this data makes a compelling case that a curved/kinked *R*_*t*_ is consistent with the population from NY.

**Figure 1.**
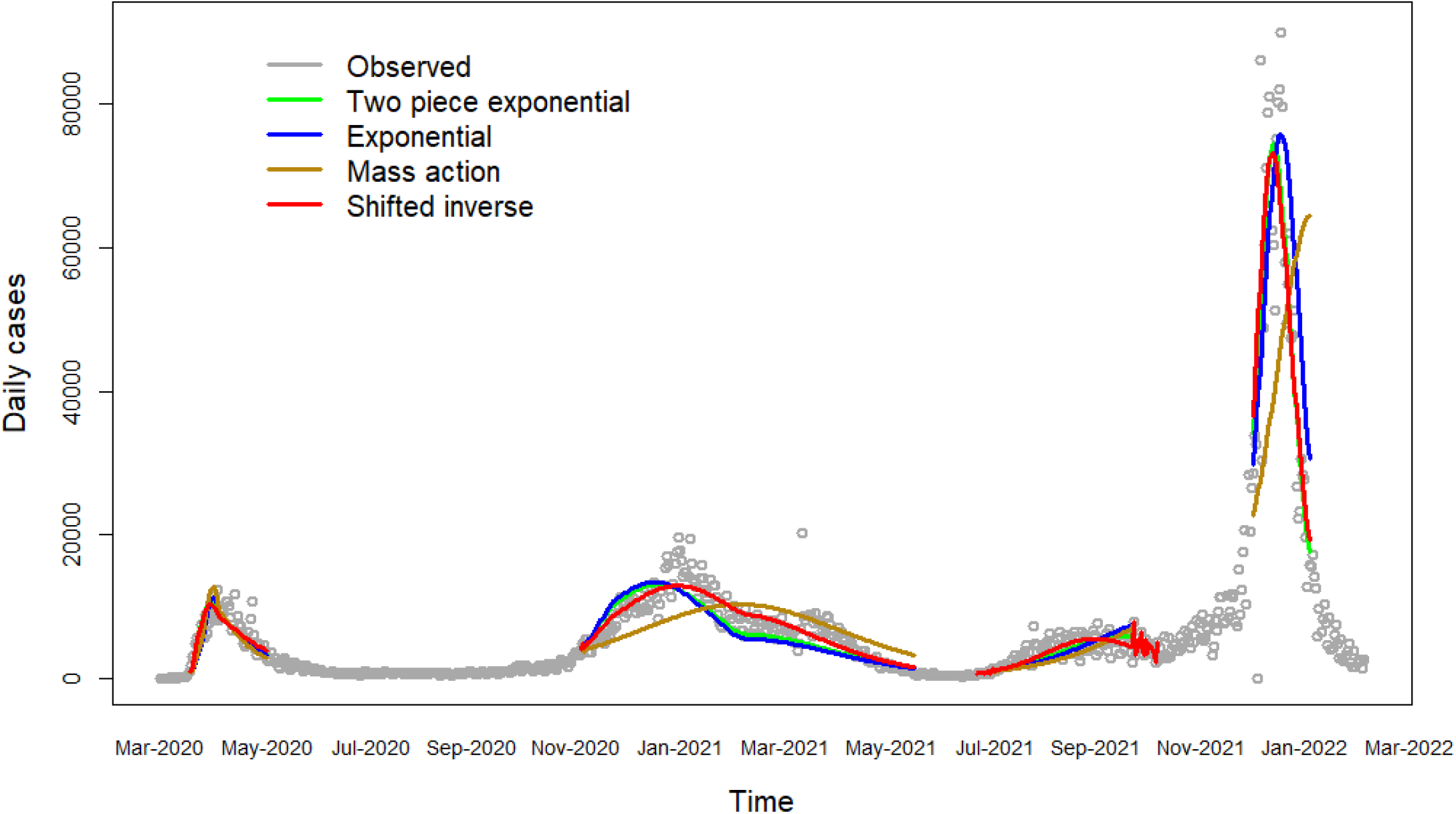
Covid-19 infection data and model fits for NY. The fits are from the beginning to the end of the waves, as defined in the text.

**Figure 2.**
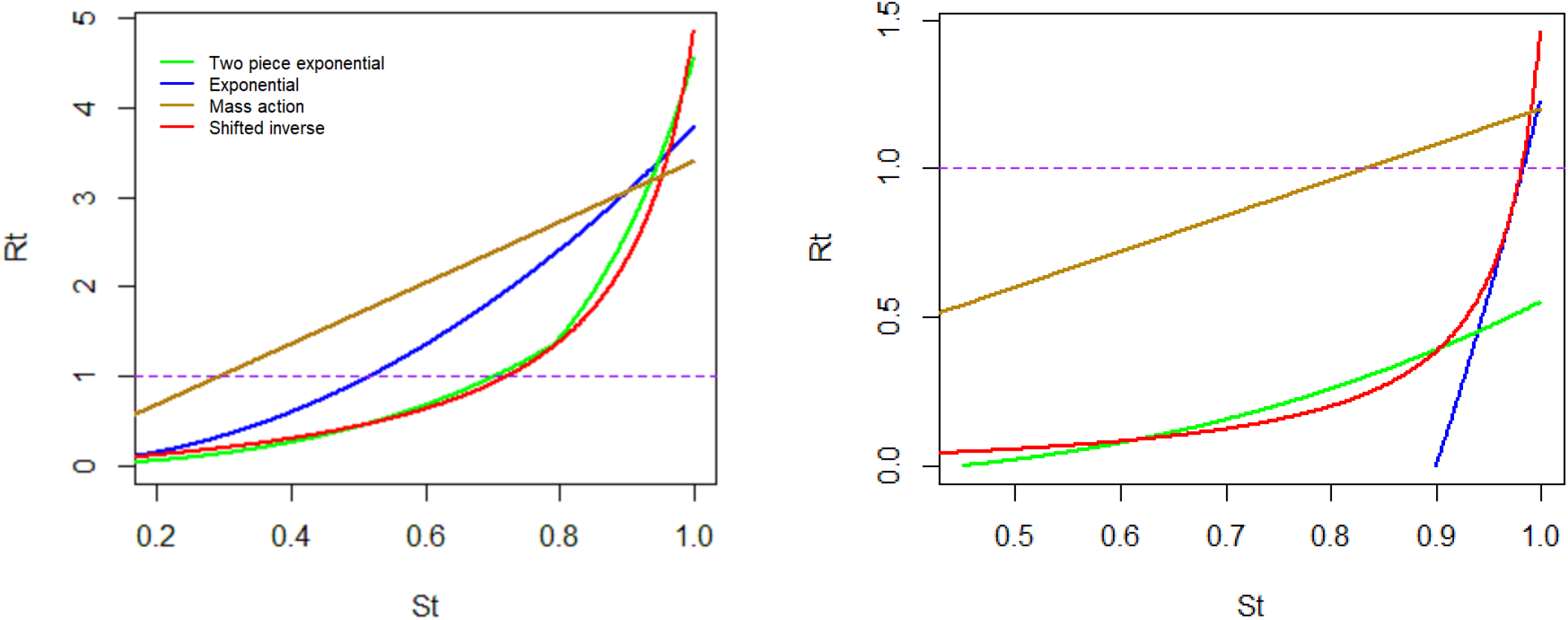
R_t_ in an unrestricted network, over the range of S_t_, as implied by the fitted models, for the NY dataset (left) and for Winnipeg (right). The endemic level (R_t_ = 1) is shown as a dashed line.

For Winnipeg, the fitted curves are given in Figure S2 in the Supplementary Material. The heterogeneous models again fit the data better than the mass action model. In this case, the best fit is given by the two piece exponential contact rate (Table S1). From Figure 2 we notice first that the initial implied *R*_*t*_ (about 1.2) is lower compared to that from New York (which is above 3 for all models). This is consistent with our expectation that a small, spread-out town will have a smaller peak *R*_*t*_ value. Still, the speed of decay is significantly greater than that implied under homogeneity, an even steeper than in NY, perhaps surprisingly. This suggests that in this case also, a flexible model for *R*_*t*_ should be used.

### 3.3 Prediction study

A prediction study is carried out for each wave in the New York dataset separately. Namely, for each day in a wave, the model is fit using all available information at the time, and the epidemic curve for the rest of the wave is predicted. Prediction quality is assessed as the mean squared prediction error of the future curve, averaged over all days. Predictions for the different shapes of *c*(·) are compared against one another and with a baseline mass action model which assumes a constant *c*(*S*_*t*_).

Results are summarized in Table 4. It is important to note that due to automation of this prediction study, fits could not be inspected visually at each time point. Thus, better fits and predictions may be possible by using different optimizer settings at each time point, and the errors presented here are likely greater than what the models would produce in actual practice.

**Table 4.** Results of prediction study for the New York data. The average represents the root of average squared deviations over all the days in the 4 waves.

| **Prediction Root MSE (in ‘000)** | **Mass action** | **One piece exponential** | **Two pieces exponential** | **Shifted inverse** |
| --- | --- | --- | --- | --- |
| **Wave 1** | 121.0 | 21.4 | 32.6 | 97.4 |
| **Wave 2** | 8.4 | 5.0 | 5.6 | 7.1 |
| **Wave 3** | 33.0 | 32.9 | 24.9 | 11.7 |
| **Wave 4** | 107.5 | 77.4 | 94.2 | 53.8 |
| **Average** | **35.3** | **17.6** | **18.1** | **25.5** |

Among the four waves, the first was least well predicted across the board, which might be due to fewer observations being available, making the models over-parameterized. All three shapes considered performed much better than the mass action model. The differences between the two exponential decay curves are relatively small, which is interesting given that the two piece model had significantly better overall fit. The lack of prediction performance might be due to the added difficulty in calibrating a model with one extra parameter. The shifted inverse model had the best prediction performance for the last two waves. This suggests two things: on one hand, that a parsimonious model will have an advantage in prediction, as seen from the fact that the shifted inverse (2 parameters) and one piece exponential (3 parameters) outperformed the more complex two piece exponential shape (4 parameters), at least one at a time. On the other hand, that choosing a well-fitting parametric model makes a stark difference in performance; here, the parametric decay models had a much better fit compared to mass action, and roughly the same ranking was preserved in terms of prediction performance.

## 3. Discussion

This paper introduced an integrated framework for prediction of infectious disease counts, based on the time-varying effective reproductive number. While an SIR model based explicitly on Rt has been formulated recently (Abbott et al., 2020), attempting to model Rt in a semi-parametric fashion, as presented here, is novel, as is demonstrating its prediction performance in a retrospective study. There are a few key arguments we make about why this Rt-based framework is preferable to the homogeneous mixing assumption, which is still used in most state-of-the-art models of infectious disease. To recapitulate:

1. The average contact rate of the infectious set, as given by curve *c*(·), is a relatively stable feature of the population which can be used to predict *R*_*t*_ at any stage of the epidemic (as measured by *S*_*t*_), and for any pathogen. Time-varying models of *R*_*t*_ do not have this feature, and they can only estimate *R*_*t*_ either retrospectively (Wu et al., 2020), which is not useful for prediction; or starting from a fully susceptible population (Chowell et al., 2016), which does not accommodate subsequent waves, unless there is full loss of immunity in the population before each new wave.
2. Formulating *R*_*t*_ as *α* × *S*_*t*_ × *c*(*S*_*t*_) is interpretable and allows for differential modeling of public health interventions by distinguishing between measures to prevent person-to-person (p2p) transmission, and measures to reduce (eliminate) social connections. These translate respectively into a reduction in *α*, and a scaling down of the argument of *c*(·) by a factor.

One of the merits of this approach is that it decouples the epidemic dynamics into a part for the pathogen (parameters *ν*; a_0_, v_2_, v_3_, etc.) and a different part for the population structure (captured by the contact rate parameters *a*_1_, …, *a*_4_), and the two are distinct up to a common scaling factor. This distinction is not obvious in other models, which often conflate the effects of p2p transmissibility and community connections. This parameterizations of *c*(·) are dependent on the population alone and could be used for predicting scenarios for new strains or pathogens, even before an outbreak begins in the community (as long as we know the relative p2p transmissibility). Another novelty is the *ψ* parameter, which specifically describes changes in the connectivity of the network, and can be used to model the effect of public health interventions designed to disrupt or reduce connections.

Another advantage of this paper is computational. Likelihood-based inference is very fast; the most complex model considered takes about one minute to fit to the full dataset. This is significantly faster than a similar Bayesian approach fit via Markov Chain Monte Carlo (MCMC). During a public health emergency when such models are needed, speed is a clear advantage.

The main limitation of this approach is that due to the large number of parameters to be estimated, the MLE search routine can get trapped in local maxima in the likelihood surface. This means that different “optimal” fits may be found when starting from different initial values. This problem is by no means unique to these models; virtually all ODE models which are fit by optimizing some loss function will encounter the same problem. One mitigation strategy is starting the optimizer from two or a few different initial values, which increases the chances of finding the global maximum. A more radical solution lies in choosing more parsimonious models, as this problem usually compounds with increasing complexity: a higher-dimensional parameter space increases the opportunity to have parameter combinations that produce similar likelihood values in different (distant) parts of the search space; this may create either ridges, or ‘bubbles’ of local maxima. While this may not be a major concern for obtaining good predictions, which tend to depend more on having a good fit rather than a realistic parameter set, caution is warranted when inferring individual parameters, and any results should be interpreted through the lens of expert opinion. Another limitation is that this model was presented in the SIRS framework, whereas COVID-19 data is often analyzed as SEIRS, which includes an intermediate exposed (E) compartment. For prediction purposes, having an exposed state is probably not essential, and we can think of the p2p transmissibility parameter *α* as the probability of transition through both states, i.e., *α* = *P*(*S* → *E*) × *P*(*E* → *I*). However, if the application requires more specialized dynamics of the exposed state to be modeled, it is entirely possible to modify the present model to accommodate this complexity, in a similar way to how ODE models currently account for this (e.g., Tang et al., 2020).

A future direction of research is relating the average contact rate *c*(*S*_*t*_) to information about the distribution of contacts in the population, obtained from either survey data (Feehan & Mahmud, 2021), or cell phone mobility logs (Chang et al., 2021). Either of these could be used to inform the degree distribution of nodes in the network (variable *K* in the Appendix) and we plan to explore this in a follow-up paper. The benefit would be twofold. First, one would know in real time how the network of connections shifts in response to public policy, instead of having to wait for data to accumulate and estimate the effect retrospectively. This would result in better predictions, as has been demonstrated by using cell phone mobility data (Chang et al., 2021). Secondly, information about the network (such as the moments of *K*) will allow an analyst to explicitly evaluate the effectiveness of public health interventions on disease spread. For instance, it will be possible to compute the *ψ* parameter for an arbitrary *x* % reduction in the average number of contacts throughout the population, and thus know exactly what impact this reduction will have on future incidence. The ability to perform calculations such as fine-tuning *x* (in this example) would be very valuable to public health officials by helping them contain a current epidemic without unduly disrupting social and economic contacts in the population.

## Data Availability

All data produced in the present study are available upon reasonable request to the authors.

https://www.gov.mb.ca/health/publichealth/surveillance/covid-19/index.html

https://github.com/nytimes/covid-19-data/blob/master/us-states.csv

## Data and code availability

All data used in this paper is publically available. We plan to release an R package at a future date, with the forecasting functionality described in the current paper.

## Appendix

### Mathematical network models

#### Definition of network quantities

Following Romanescu & Deardon (2017), assume that the population can be described by a fixed network with individuals as nodes, and connections as edges, and define the degree of a node (number of emanating edges) via random variable *K* with distribution *P*(*K* = *k*) = *p*_*k*_, *k* ≥ 0. Then, a basic result is that the chance of an individual becoming infected is proportional to its degree; explicitly, the survival probability of a susceptible with degree *k* at some time *t* is 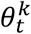, for all *k* ≥ 0 (Volz, 2008; Miller, 2011). Furthermore, *θ*_*t*_ is linked to the susceptible fraction via *g*(*θ*_*t*_) = *S*_*t*_, where *g* is the probability generating function, or pgf of *K*, namely 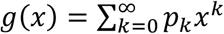. To describe transmission dynamics, it is of interest to obtain the distributions of 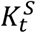 and 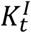, the degrees of the susceptible and the infectious sets at time *t*. Their pgf’s are 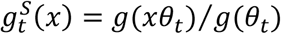, and 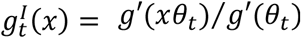, respectively.

#### Dependence of R_t_ on the network

The definitions above allow us to derive the mean and variance of the expansion factor *X*_*t*_, which is the number of secondary infections for one infected individual. Since 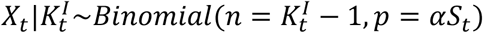, where *α* is the per-edge probability of transmission over the entire infectious period, we arrive at

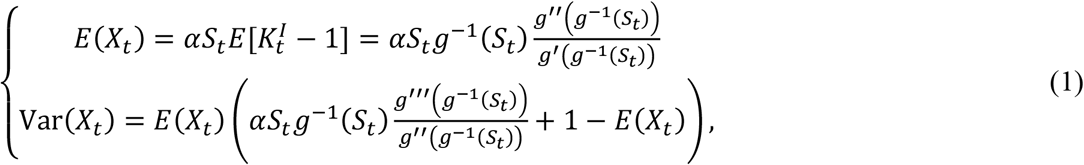

where *g*^−1^ is the inverse function to *g*, and *g*′, *g*″ and *g*‴ are the 1^st^, 2^nd^, and 3^rd^ derivatives of *g*. Importantly, *E*(*X*_*t*_) = *R*_*t*_, the effective reproductive number.

As we are not likely to observe *p*_*k*_ or *g* directly, the take-aways are firstly that *R*_*t*_ = *αS*_*t*_*c*(*S*_*t*_), where *c*() is the average contact rate of the infectious set (less one). Secondly, that *c* is a function of the network that evolves in time via *S*_*t*_, in other words *c*(*t, S*_*t*_) = *c*(*S*_*t*_), provided that the structure of the network doesn’t change. Thus, *S*_*t*_ emerges as the natural “clock” of the epidemic. As a note on language, we refer to *c* as the average contact rate of the infectious set for shorthand, but, in fact, it should be made clear that 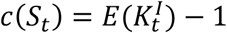, which reflects the fact that transmission can proceed to any of the connections except for the original infector.

#### Common network distributions

Three degree distributions found in the literature include:

- The Poisson distribution, 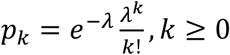. The pgf is *g*(*x*) = *e*^*λ*(*x*−1)^, and *R* = *αS* (*λ* + *ln*(*S*_*t*_)).
- The geometric distribution: 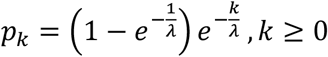, had pgf 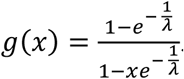. *R*_*t*_ simplifies to 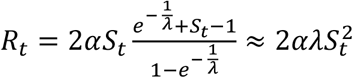, for large *λ*.
- The power law with an upper cutoff has *p*_*k*_ ∝ *k*^−*λ*^, for *k* = 1,2, …, *k*_*max*_. Here, the pgf is 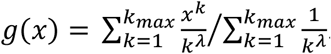. In this case there is no closed form solution for *R*_t_, however, the power law has the heaviest tail of the three distributions, which leads *R*_*t*_ to peak the highest (see Fig 1 in Romanescu & Deardon (2017) for an example of a simulated epidemic and the corresponding *R*_*t*_).

As can be seen in these common degree distributions, the function *f* is increasing, which makes the decay of *R*_*t*_ faster than under the mass action assumption.

### **Derivation of** 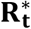

First, it is straightforward to show that weighting the probabilities *p*_*k*_ by an exponential tail results in the new probabilities 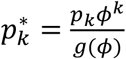.

We find *g*∗ (*x*) as follows:

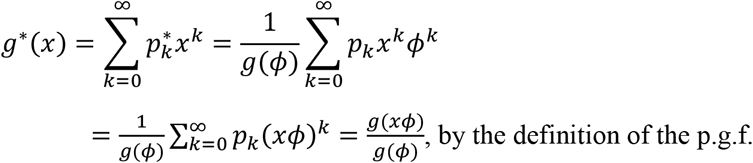

To find *g*∗^−1^(*x*), let *x* = *g*∗ (*y*) and solve for *y*:

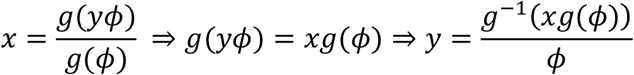

Therefore, 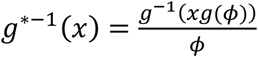. Finally,

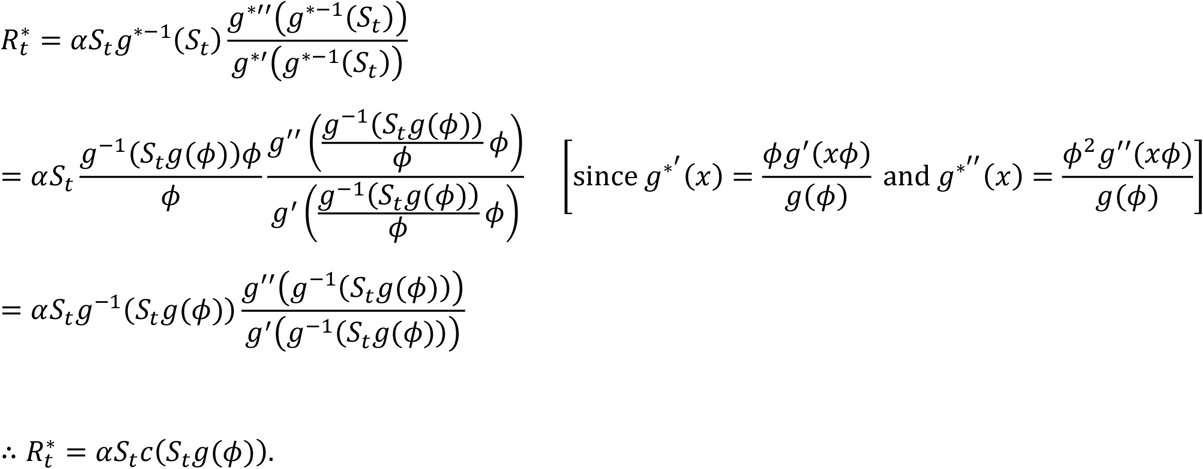

### Estimators of the effective reproduction number

#### Instantaneous reproduction number

This backward-looking estimator was proposed in Cori et al. (2013), though it can be found earlier in Nishiura & Chowell (2009). It compares the number of new infections on day *t* with the infections prior to *t*. The mathematical formulation of the estimator is 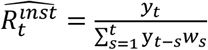, where *y*_*t*_ is the number of infections on day *t* and *w*_*s*_ is the generation interval. This method requires a distributional assumption about the form of generation interval.

### Justification of SIRS assumptions

The following derivation is based on the one in Romanescu & Deardon (2017). If *p*_*k,t*_ is the probability that a random susceptible at time t has degree *k*, and the corresponding p.g.f. is *g*_*t*_, then at time *T*_*l*_ we have 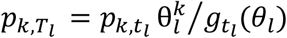. Also, since 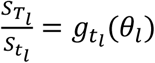, we obtain 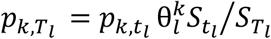. The balance equation at time *t*_*l*+1_ for susceptible nodes of degree k is:

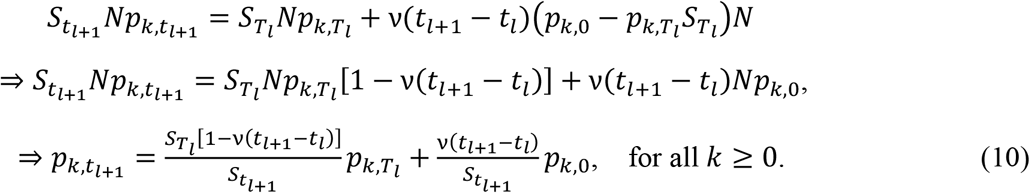

Equation (10) implies that, at the beginning of wave *l* + 1, the degree distribution of susceptible nodes is a mixture of distributions 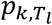 and *p*_*k*,0_ with weights 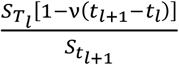 and 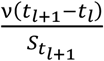. (It can be verified that these sum to one.) Another way to say this is that the susceptible population network at time *t*_*l*+1_ can be partitioned into two subnetworks of sizes 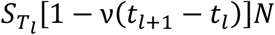 and *v*(*t*_*l*+1_ − *t*_*l*_)*N* with degree distributions 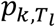 and *p*_*k*,0_, respectively.

To derive the spread dynamics in the two subnetworks, start at time *t*_2_ (assuming a first wave has passed through the fully susceptible population at time *t*_1_). Infectious individuals in the first subnetwork will have a contact rate that decreases from the level it was at *T*_*l*_, as nothing has changed for this group; this is given by 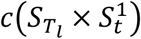, where 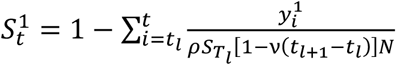 is the fraction of susceptibles in this subnetwork (starting from 1 at time *t*_2_). Similarly, the subnetwork having original degree distribution will have a contact rate of the infectious set that evolves as 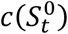, where 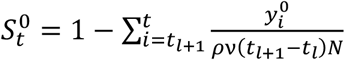, i.e., as in a fully susceptible population. The total predicted number of cases aggregates the contributions of each subnetwork:

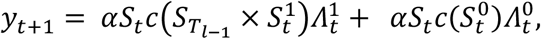

where 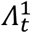 and 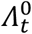 follow the same notation. Note that because the original network was built according to the Configuration Model (Molloy & Reed, 1998), there is no preference for an infected node to pass the infection to susceptibles in its own subnetwork. In fact, new cases (*y*_*t*+1_) should be split according to the total number of edges remaining in each susceptible subnetwork. Since we don’t know the exact degree distribution, we will use the number of susceptible nodes in each subnetwork as a proxy to determine the split between 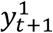 and 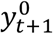, namely

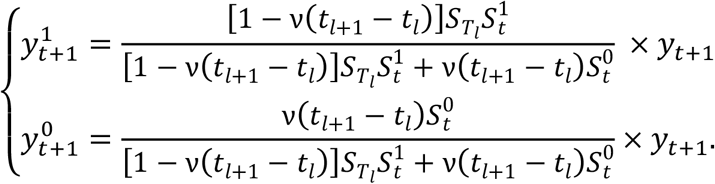

For years 3+ we will assume the same formulas as above for *l* = 3,4, …, namely that the two subnetworks have contact rates 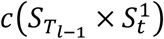 and 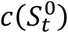 for times *t* during wave *l* (*t*_*l*_ < *t* < *T*_*l*_). An exact solution exists (see Romanescu & Deardon, 2017; Bansal et al., 2010), however it relies on knowledge of the original degree distribution.

## Supplementary material

**Figure S1.**
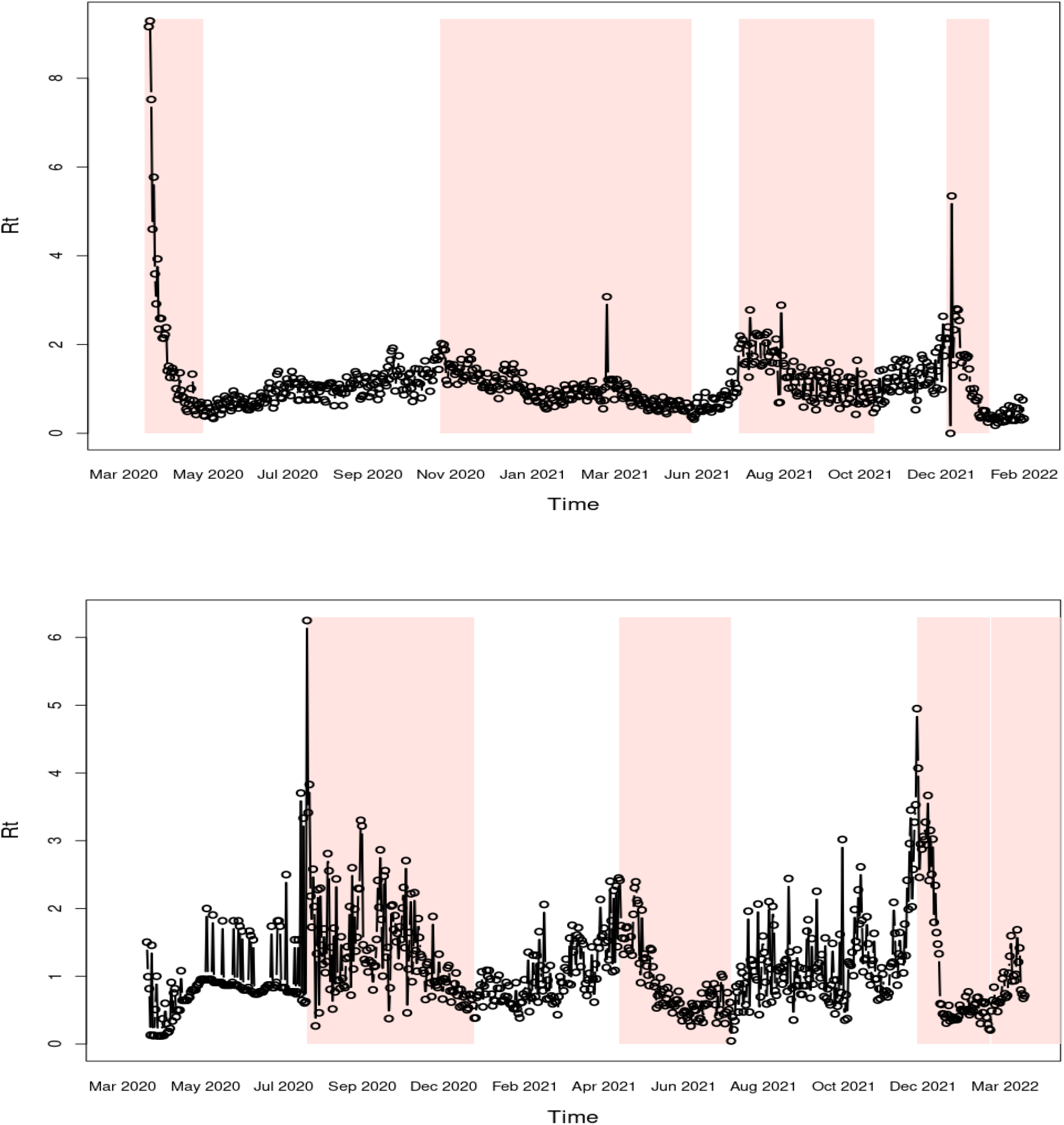
Plot of the empirical Rt used to define the beginning and end of epidemic waves. The waves are shown as shaded regions for the New York (top) and Winnipeg dataset (bottom). We use a uniform serial interval over 20 days for computing Rt, as this produces a smoother estimate and enhanced visibility of patterns, compared to the log-normal serial interval.

**Table S1.** Parameter estimates for the Winnipeg RHA, using the full dataset. Maximum likelihood estimates and their standard deviations are shown. The (negative) log-likelihood value and the AIC are given at the bottom.

| Parameter |  | Definition | two-piece<br>exponential | one piece<br>exponential | shifted<br>inverse | mass<br>action |
| --- | --- | --- | --- | --- | --- | --- |
| $\alpha \times c(S_t)$ | $a_0$ | linear multiplier | – | 11.20<br>(0.15) | 0.06<br>(0.002) | 1.20<br>(0.008) |
| | $a_1$ | speed of decay | 0.80<br>(0.41) | 1.05<br>(0.009) | 1.04<br>(0.002) | – |
| | $a_2$ | final size | 0.45<br>(0.25) | 0.90<br>(0.001) | – | – |
| | $a_3$ | speed of decay (early) | 11.11<br>(0.457) | – | – | – |
| | $a_4$ | transition point/shift | 0.95 (–) | – | – | – |
| $\nu$ | | loss of immunity rate | 1.64 (–) | 0.70<br>(0.046) | 1.53<br>(–) | 0.00<br>(0.043) |
| relative variant<br>transmissibility<br>($v_1 = 1$) | $v_2$ | for second wave | 1.01<br>(0.018) | 1.42<br>(0.018) | 0.98<br>(0.018) | 0.95<br>(0.022) |
| | $v_3$ | for third wave | 1.53<br>(0.039) | 2.29<br>(0.087) | 1.43<br>(0.032) | 0.94<br>(0.013) |
| restriction<br>factors ($\psi$) | $\psi_1$ | limited reopening | 1.00 (–) | 1.00 (–) | 1.00 (–) | 0.64 (–) |
| $u$ | | overdispersion factor | 27.78<br>(2.363) | 40.55<br>(3.46) | 43.85<br>(3.86) | 235.41<br>(27.013) |
| $-l(\theta)$ | | neg. log likelihood | 1587.806 | 1645.23 | 1656.995 | 1830.46 |
| # of parameters |  |  | 9 | 8 | 7 | 6 |
| AIC |  | Akaike Information<br>Criterion | 3193.6 | 3306.5 | 3328.0 | 3672.9 |

**Figure S2.**
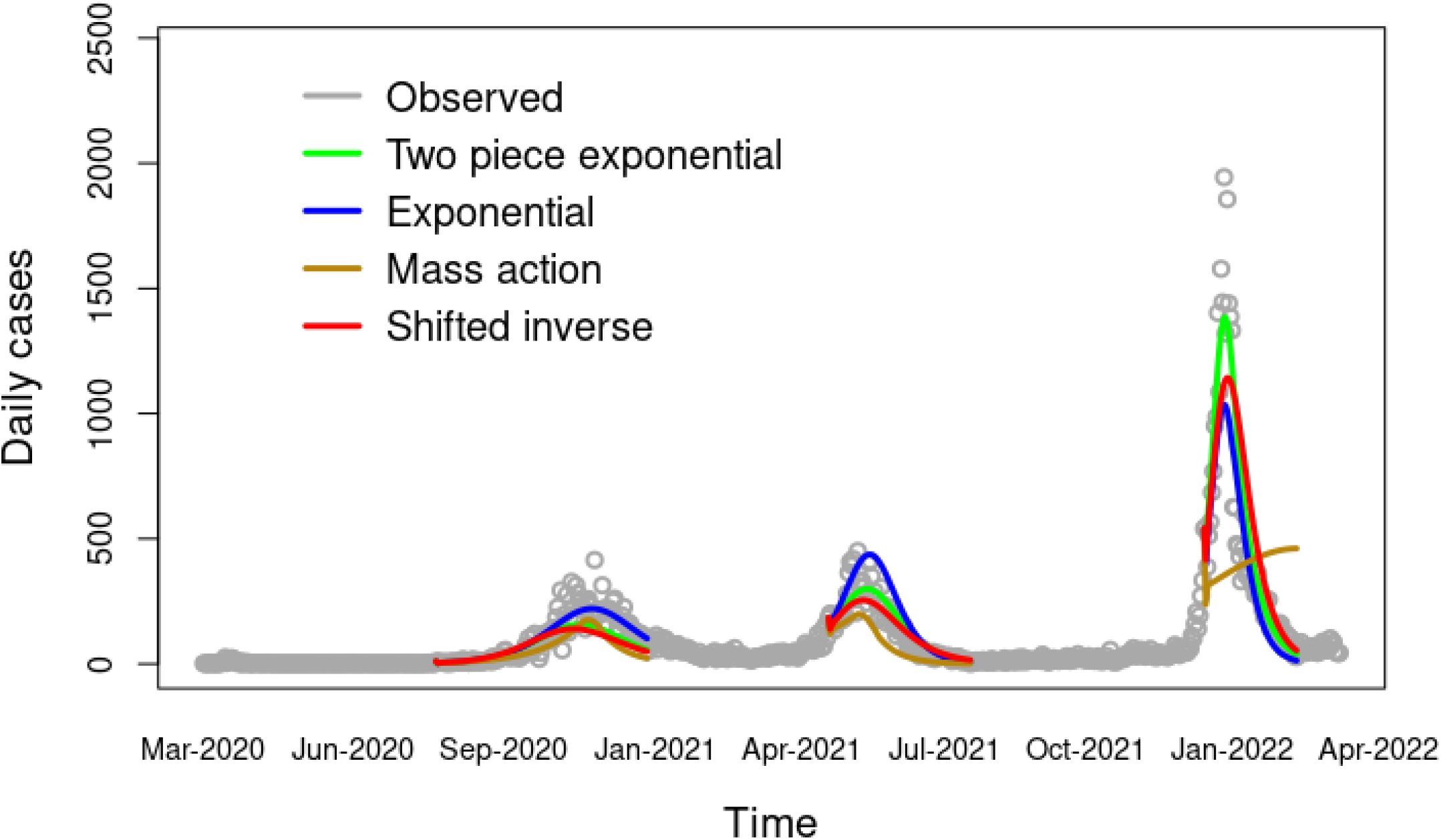
Covid-19 infection data and model fits for NY. The fits are from the beginning to the end of the waves, as defined in the text.

